# Screen Time and Musculoskeletal Neck Pain in Children: A Comprehensive Systematic Review and Lifestyle Recommendations

**DOI:** 10.1101/2024.04.28.24306242

**Authors:** Huzaifa T Rampurawala, Samir H Rampurawala

## Abstract

Musculoskeletal neck pain is one of the leading ailments in the world right now and affects everyone, from seniors to prepubescent kids. Neck pain also costs countries billions of dollars in healthcare and medical expenses. Research in this field is slim, and thus, a compilation of this information is necessary. This systematic review aims to collect articles worldwide, exploring the correlation between screen use and neck pain in children. This systematic review harnesses PubMed, JSTOR, and Google Scholar data to comprehensively analyze 6,804 articles on the subject, cutting it down to 13 papers. To do this, an independent reviewer first distinguished note-worthy articles from said databases and used articles that fit this study. Then, the articles with data that fit the variables were used. Preliminary results in all articles indicate a substantial positive correlation between reduced screen time and reduced instances of neck pain issues, signifying the potential for lifestyle changes in children and adolescents. This systematic review also highlights its recommendations for screen use at different points in a child’s life, allowing parents to determine their kids’ best screen use rate. By synthesizing these findings, this review offers valuable insights into the potential benefits of reducing screen time as a preventive measure against neck pain in adolescents, increasing support for this cause, and expanding informed parental guidance in managing children’s screen usage habits. These recommendations were determined based on data from articles in the systematic review. Additional work in this field focusing on screen use and neck pain in adolescence is needed, and a higher-quality recommendation chart must be manufactured.

## Introduction

Musculoskeletal pain is a significant problem and affects both adults and children. In 2016, treatments for low back and neck pain had the most significant healthcare expenditure in the United States, with an estimated 134.5 billion dollars being used for healthcare spending and other musculoskeletal disorders coming in second.^1^ With an association with long-term sickness and early retirement^2^, low back and neck pain have caused a significant socioeconomic burden. The World Health Organization (WHO) ranked neck pain and other musculoskeletal diseases at 4th and 10th, respectively, among health conditions with years lived with a disability.^3^ Musculoskeletal pain is also highly prevalent in younger individuals, affecting up to 40% of adolescents.^4,5^ Neck pain is also ranked seventh for most years lived with a disability based on the WHO Global Burden of Disease for 15-19 year olds^6^, and studies have linked musculoskeletal pain in adolescence to musculoskeletal pain in adults.^7–9^ Nonetheless, neck pain in adolescents has not been as extensively studied as in adults.

Screen use can induce neck and back pain, eye strain, and shoulder and arm pain. It can also lead to psychological and social effects^10^. Changes in lumbar lordosis and thoracolumbar kyphosis are also linked to excessive screen use and lower back pain.^11^ Screen-based activities and sedentary lifestyles contribute to upper quadrant musculoskeletal pain (UQMP) in adolescents^12^. Studies show that young adults are also influenced by screen time use, with a correlation between mobile phone usage and neck pain in university students^13,14^; however, some studies have found no or little correlation between screen usage and neck pain. Diana et al. found little correlation between TV, computer, and video game usage and neck pain in school-aged children, and Pirnes et al. had mixed results on screen usage and neck pain in children.^15,16^ This shows that while evidence of a correlation between screen use and human anatomy has been found, more work is needed to provide a prominent statement for adults and kids. This systematic literature review and meta-analysis will analyze evidence to understand the relationship between neck pain and screen usage and provide a timeline for parents and children so they can use the optimal amount of screen time for their necks.

## Methods

### Search Strategy

We carried out this study following the PRISMA 2020 statement. This study used searches of PubMed, JSTOR, and Google Scholar for records through July 30, 2023. Based on our research question, the search strings focused on neck pain and screen use in adolescents and children. The strings used are available here.

### Eligibility Criteria

For the pre-screening process, we removed duplicate articles using Mendeley and removed search terms, increasing the number of irrelevant papers. After eliminating duplicates, one independent reviewer screened and reviewed the title and abstract, gathering relevant documents to our research question. The reviewer then reviewed the complete text, removing papers that omitted important information (age range, pain assessment, data collection methods).

### Inclusion Criteria

We examined all observational studies (cross-sectional, case-control, or longitudinal designs) associated with age (specifically adolescence), neck pain, and screen use. We investigated all studies associated with these criteria without a restriction on race, gender, or publication date.

### Data Extraction

An independent reviewer extracted data from the included articles. The data included the name, age range, sex ratio, type of study, sample size, neck pain data assessment, type of data collection assessment, location, and the study results. Once the reviewers fully extracted data from the studies, the information was reviewed to add details to the data.

### Quality Assessment

An independent reviewer went through the National Institute of Health (NIH) quality assessment tool to perform a quality assessment. This tool is a 14-item assessment of the quality of each article. It is used for cross-sectional and cohort studies, qualifying the case as good, fair, or poor.

### Timeline

One reviewer reviewed the different sources to create a timeline and listed the inappropriate screen use limits associated with each age. After compiling these studies, the reviewer incorporated each paper’s recommended screen use, age, and odds ratio. Using a weighted bi-exponential interpolation method, we went through each age and determined the odds ratio between each age. After that, the odds ratio was grouped into different intensities (1-10). While this limit should be followed, appropriate leniency and consideration should be used towards time spent without a break, movement while watching, and entertainment and work-related activities.

### Study Selection

Using the search strings mentioned above, 6,804 records were found, 5,577 marked as ineligible, and 24 duplicates, bringing the total to 1203 records screened. Of these, 1146 articles were excluded based solely on the title and abstract, and 18 reports were not included due to the cost of accessing the article. Thirty-nine reports were investigated further via full text. Figure 1 shows the full PRISMA flow chart.

## Results

### Study Characteristics

Of the 8,804 articles collected, screened, and investigated, we used 13 articles in the systematic review. During the study analysis, the participants were school and university students (6 - 25 years old) with either an equal or adjusted male-to-female ratio. Of the included research, two articles were from Brazil^18,19^, two from Portugal^20,,21^, two from Thailand^22^, and one study each was from Ethiopia^23^, Jordan^24^, Iran^25^, Shanghai^26^, Kuwait^27^, Turkey^28^, Lebanon^29^, and Taiwan^30^.

These studies had 14,353 total participants, dating from 2013 to 2023. The most extensive study was in Shanghai^26^ with 3,016 participants, and the most minor study was in Rio de Janeiro, Brazil^18^, with 150 participants. These studies collected data through interviews, questionnaires, singular questions, and scales, but the most prominent of these scales was the Nordic questionnaire or an adapted version of the Nordic Questionnaire.^31^

All studies reviewed focused on total screen use, with all subjects using mobile devices; however, several studies found different screen use options, such as tablet^19,21,22,23,25,27,28^, video games^20,21,26,28^, computers^20,21,23,25,26,27,28^, and television use^20,21,25,26,28^. According to our search results, we divided the measurable outcomes into total screen use (including total phone, tablet, video games, computer, and television use) and phone use, and we created timelines for each outcome using the available data.

**Table 1.**
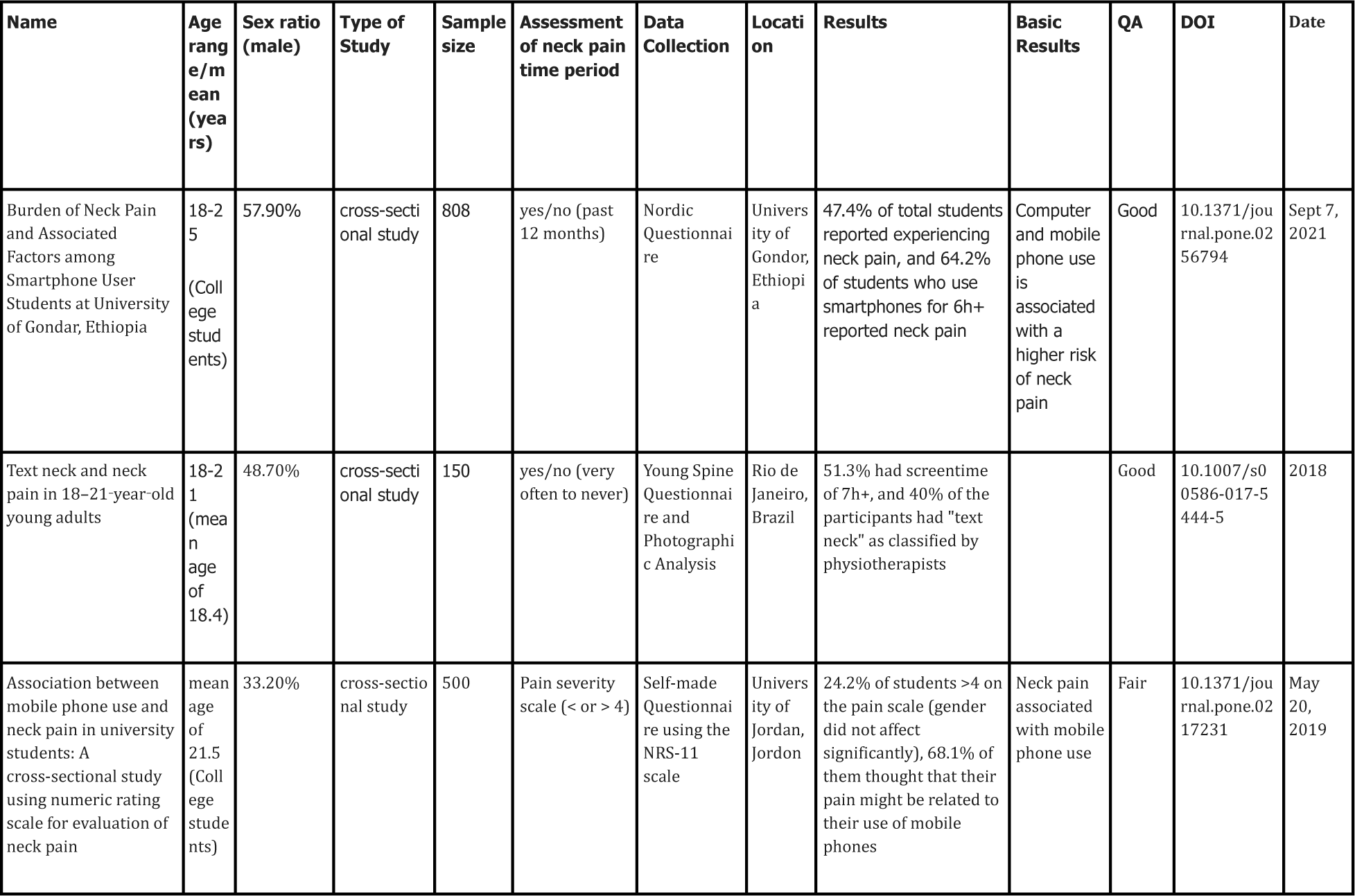
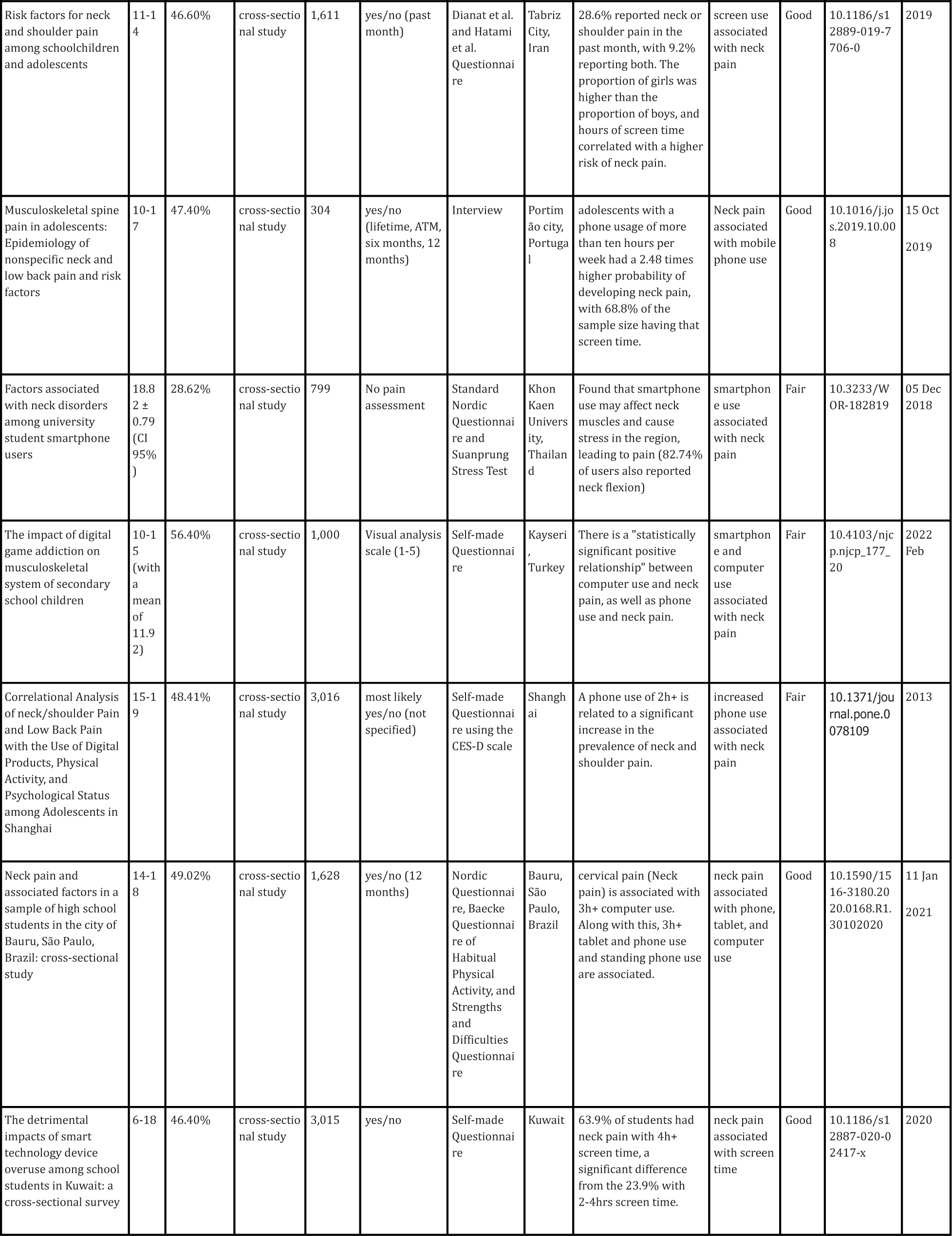
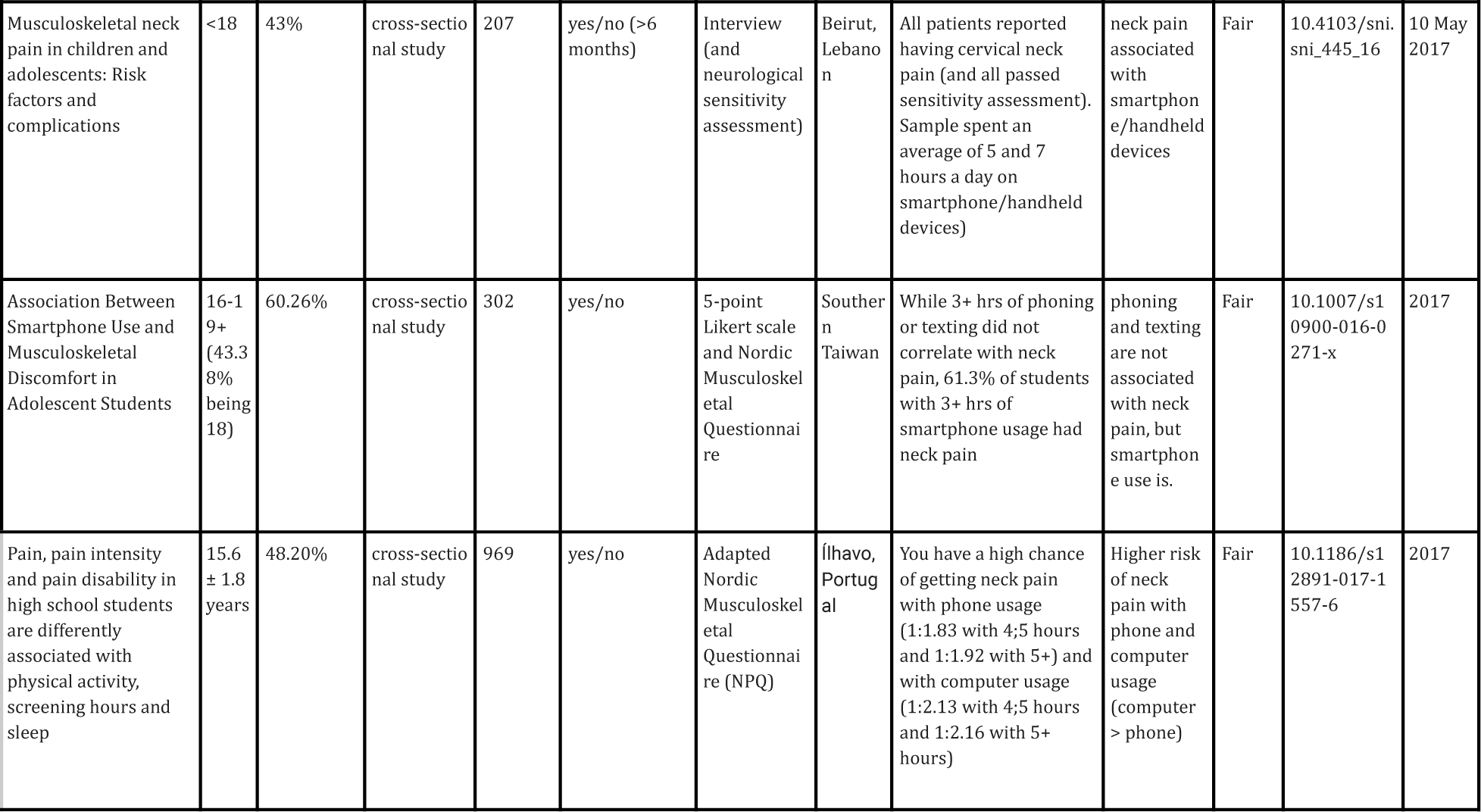
Study Information. A review of information from each study. This table showcases the extraction of data during this study. The most critical pieces of data are the age range and the results, which explain the findings; most papers correlated higher odds ratios to neck pain risk depending on age. Data collection methods were extracted to observe the type of data extraction these studies completed (direct, semi-direct, indirect). The date, sample size, and sex ratio were extracted for the quality assessment.

**Table 2.**
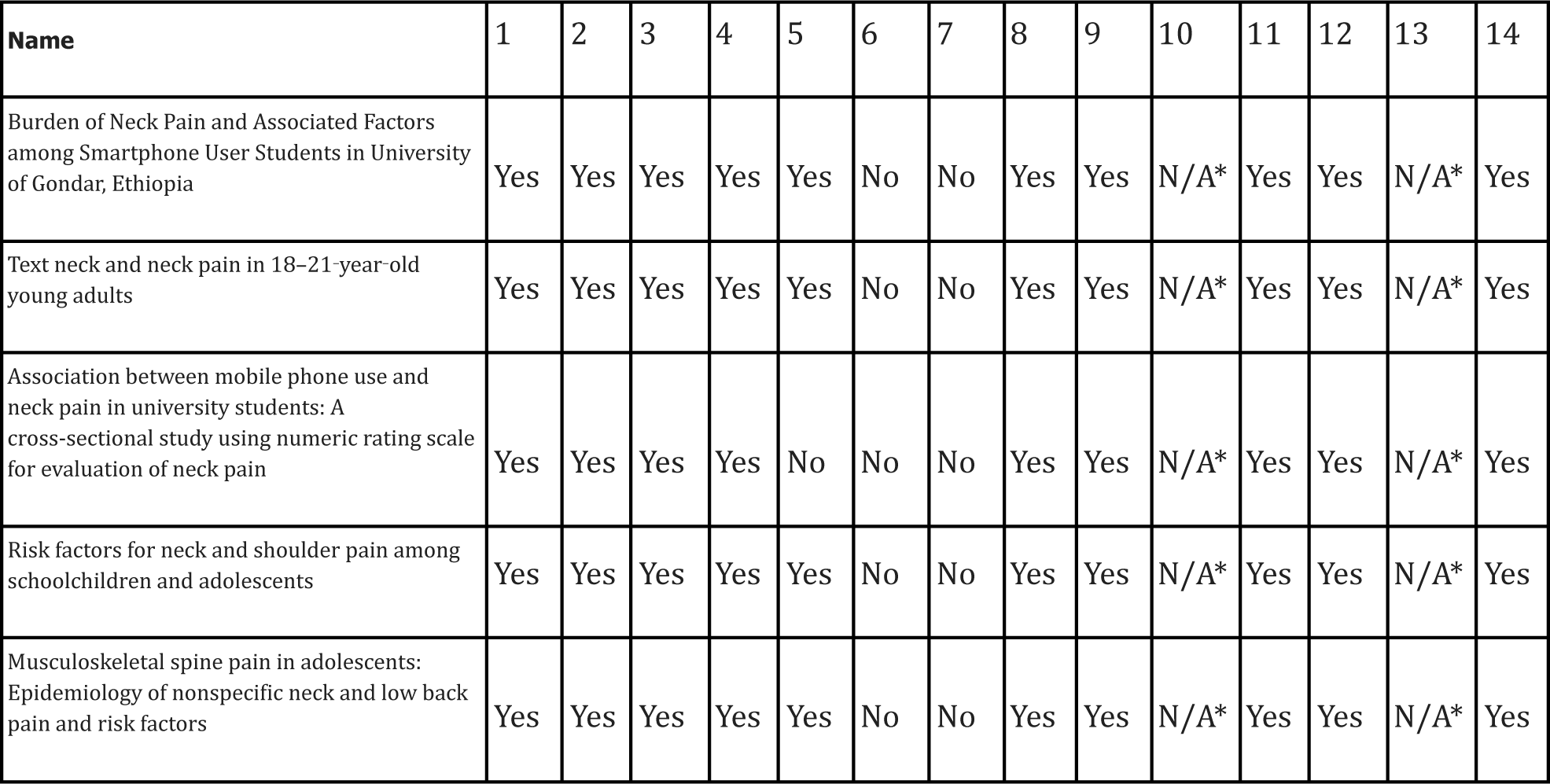
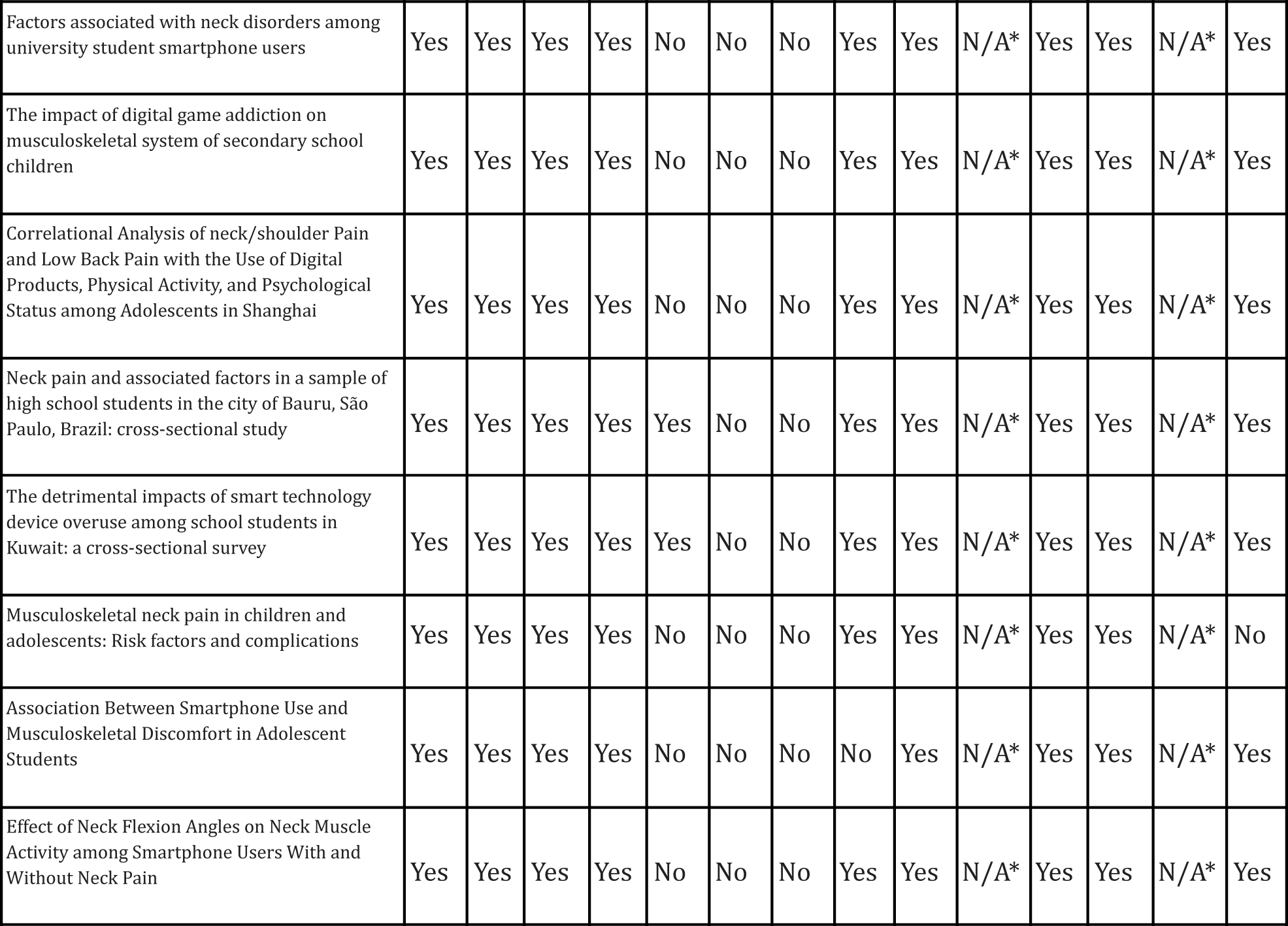
Quality Assessment. The Quality Assessment for each study was reviewed. All papers received a Good or Fair score for the questions using the NIH tool during the quality assessment. This means these articles are acceptable for this study.

| Name | 1 | 2 | 3 | 4 | 5 | 6 | 7 | 8 | 9 | 10 | 11 | 12 | 13 | 14 |
| --- | --- | --- | --- | --- | --- | --- | --- | --- | --- | --- | --- | --- | --- | --- |
| Burden of Neck Pain and Associated Factors among Smartphone User Students in University of Gondar, Ethiopia | Yes | Yes | Yes | Yes | Yes | No | No | Yes | Yes | N/A* | Yes | Yes | N/A* | Yes |
| Text neck and neck pain in 18–21-year-old young adults | Yes | Yes | Yes | Yes | Yes | No | No | Yes | Yes | N/A* | Yes | Yes | N/A* | Yes |
| Association between mobile phone use and neck pain in university students: A cross-sectional study using numeric rating scale for evaluation of neck pain | Yes | Yes | Yes | Yes | No | No | No | Yes | Yes | N/A* | Yes | Yes | N/A* | Yes |
| Risk factors for neck and shoulder pain among schoolchildren and adolescents | Yes | Yes | Yes | Yes | Yes | No | No | Yes | Yes | N/A* | Yes | Yes | N/A* | Yes |
| Musculoskeletal spine pain in adolescents: Epidemiology of nonspecific neck and low back pain and risk factors | Yes | Yes | Yes | Yes | Yes | No | No | Yes | Yes | N/A* | Yes | Yes | N/A* | Yes |
| Factors associated with neck disorders among university student smartphone users | Yes | Yes | Yes | Yes | No | No | No | Yes | Yes | N/A* | Yes | Yes | N/A* | Yes |
| The impact of digital game addiction on musculoskeletal system of secondary school children | Yes | Yes | Yes | Yes | No | No | No | Yes | Yes | N/A* | Yes | Yes | N/A* | Yes |
| Correlational Analysis of neck/shoulder Pain and Low Back Pain with the Use of Digital Products, Physical Activity, and Psychological Status among Adolescents in Shanghai | Yes | Yes | Yes | Yes | No | No | No | Yes | Yes | N/A* | Yes | Yes | N/A* | Yes |
| Neck pain and associated factors in a sample of high school students in the city of Bauru, São Paulo, Brazil: cross-sectional study | Yes | Yes | Yes | Yes | Yes | No | No | Yes | Yes | N/A* | Yes | Yes | N/A* | Yes |
| The detrimental impacts of smart technology device overuse among school students in Kuwait: a cross-sectional survey | Yes | Yes | Yes | Yes | Yes | No | No | Yes | Yes | N/A* | Yes | Yes | N/A* | Yes |
| Musculoskeletal neck pain in children and adolescents: Risk factors and complications | Yes | Yes | Yes | Yes | No | No | No | Yes | Yes | N/A* | Yes | Yes | N/A* | No |
| Association Between Smartphone Use and Musculoskeletal Discomfort in Adolescent Students | Yes | Yes | Yes | Yes | No | No | No | No | Yes | N/A* | Yes | Yes | N/A* | Yes |
| Effect of Neck Flexion Angles on Neck Muscle Activity among Smartphone Users With and Without Neck Pain | Yes | Yes | Yes | Yes | No | No | No | Yes | Yes | N/A* | Yes | Yes | N/A* | Yes |

## Discussion

This review focused on the relationship between neck pain and screen use in different age groups. Studies found a positive, significant relationship between neck pain and screen use from the ages of 10 to 25.

This article opens up space for a more significant discussion of on-screen use’s impact on the development of adolescents and the anatomical problems that can stem from improper posture. Table 3 showcases this: as technology has a more significant impact on people’s lives, we slowly forget the risk it can have on kids and young adults, with the pandemic increasing the average screen time of a sample size of 228 from 4.4 hours to 6.15 hours in children age 7^32^. Studies also show that screen use has become more popular at work and school^33^ among adults and children, most likely due to COVID. This can be a huge problem for not just the mental but also the physical development of kids. The CDC attests to this, explaining that in 2018, kids ages 8-18 had an average screen time of 7.5 hours; however, 4.5 hours were spent in front of a Tv^34^. Comparing this to Table 3, you can see that the average screen time per day does not match the recommendations, which could harm future generations’ anatomical growth.

**Table 3.**
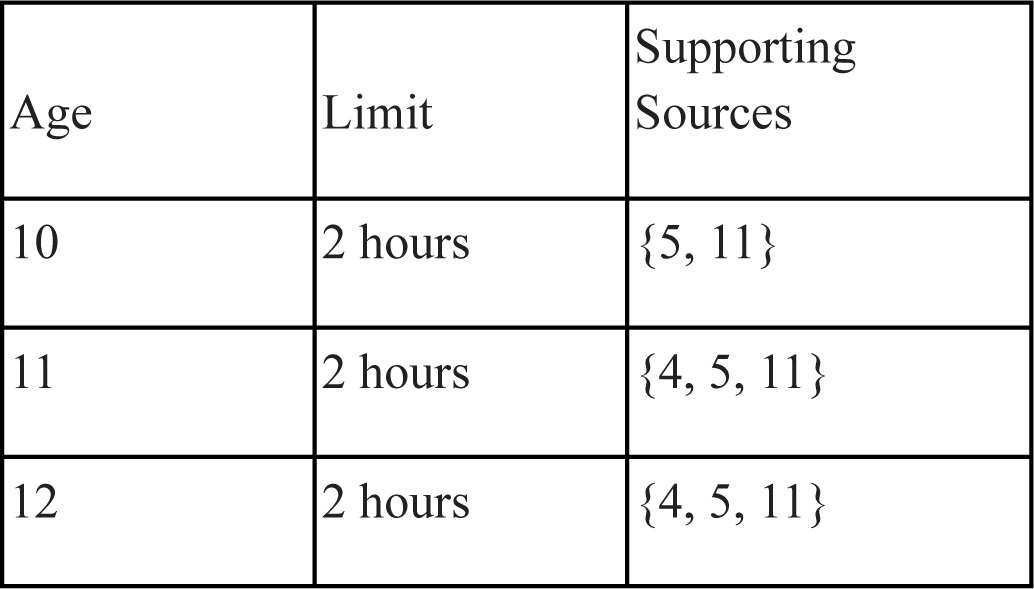
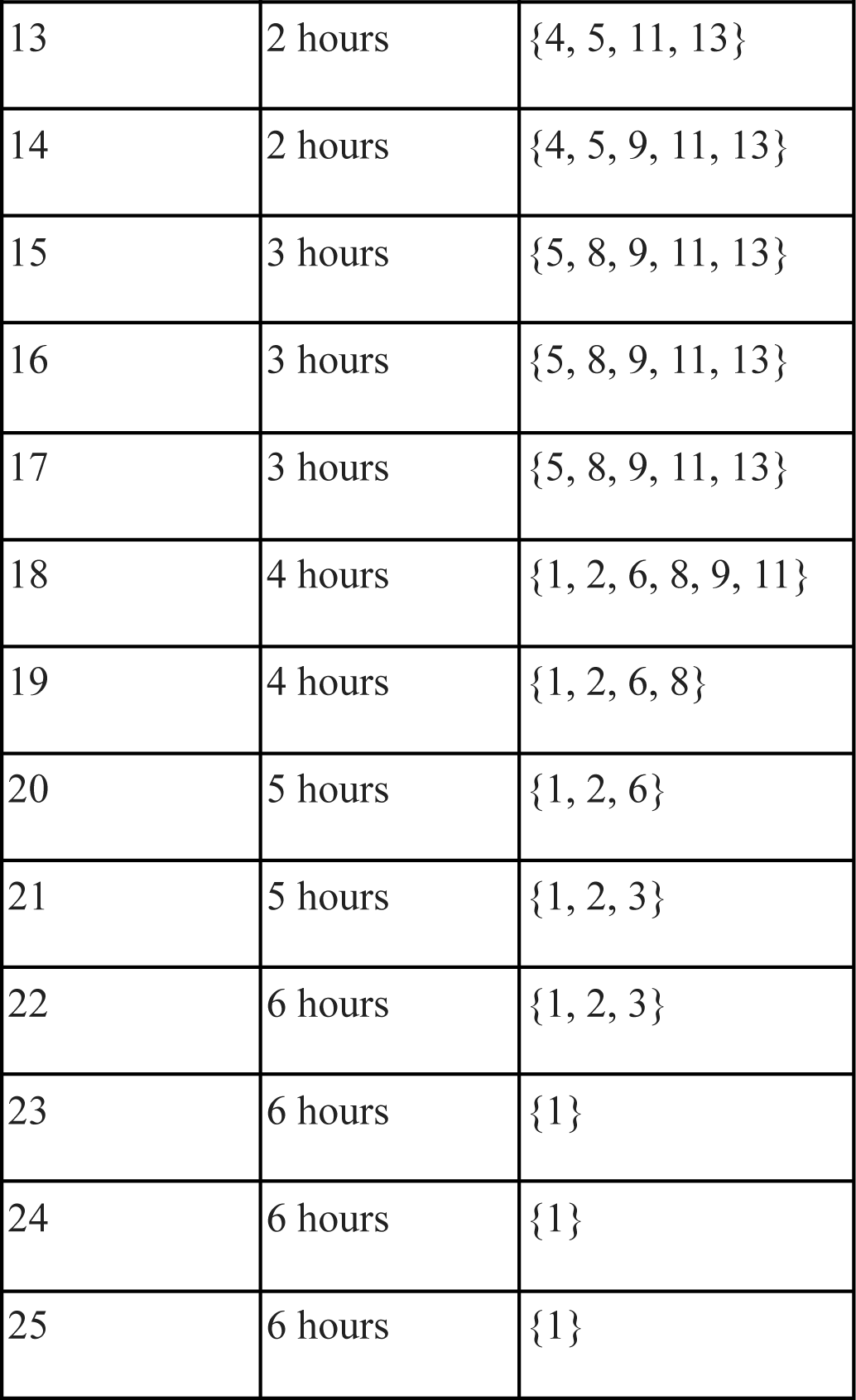
Recommendation for screen time based on age. The timeline results were built based on the sources from the systematic review. This table showcases the recommended screen time limit per age using the supporting sources. Each age also includes articles that support the reasoning behind that limit. These limits can only be used as a recommendation, as other factors can apply.

| Age | Limit | Supporting Sources |
| --- | --- | --- |
| 10 | 2 hours | {5, 11} |
| 11 | 2 hours | {4, 5, 11} |
| 12 | 2 hours | {4, 5, 11} |
| 13 | 2 hours | {4, 5, 11, 13} |
| 14 | 2 hours | {4, 5, 9, 11, 13} |
| 15 | 3 hours | {5, 8, 9, 11, 13} |
| 16 | 3 hours | {5, 8, 9, 11, 13} |
| 17 | 3 hours | {5, 8, 9, 11, 13} |
| 18 | 4 hours | {1, 2, 6, 8, 9, 11} |
| 19 | 4 hours | {1, 2, 6, 8} |
| 20 | 5 hours | {1, 2, 6} |
| 21 | 5 hours | {1, 2, 3} |
| 22 | 6 hours | {1, 2, 3} |
| 23 | 6 hours | {1} |
| 24 | 6 hours | {1} |
| 25 | 6 hours | {1} |

All studies in this review found an association between phone use and neck pain. This dynamic between neck pain and screen use could also be necessary as technology and screen use become more prevalent. Screen use is also associated with several health complications, such as obesity^35^, high blood pressure^36^, eye strain^37^, sleep problems^38^, depression, and anxiety^39^.

While the literature on this subject was thorough, the variety of neck pain screenings and questionnaires worldwide was inappreciable. While neck pain is a prevalent problem worldwide, studies and research in musculoskeletal pain don’t have extensive literature available. The available studies were of sufficient quality and worked well for the study, but most of the literature focused on students between the ages of 10 and 25. It would be exciting to review more elementary school-focused studies, though that can be challenging in the research field.

There is a significant lack of awareness and education about the overuse of screens and the associated problem of neck pain among students and teachers. The prevalence of neck pain is surprisingly high, affecting a substantial portion of the sample sizes in this review, yet this correlation has remained unchanged in the last ten years. Many studies highlight the need for a reduction in screen use, and this prevalence will only increase in the coming years.

This study has contributed significantly to understanding the relationship between neck pain and screen use. It has provided a rough timeline that can be utilized by students and parents to assess the potential risk of neck pain associated with screen use. This timeline is a valuable tool as it offers parents an estimate of the appropriate amount of screen time for their children while also allowing for some flexibility. The creation of this timeline was informed by the articles included in the systematic review, indicating that it is based on a comprehensive analysis of existing research in this area. Acknowledging that this timeline is not static and may evolve as more data becomes available is essential. Therefore, it is important to continue gathering information and updating the timeline to ensure accuracy and relevance. Developing a more robust timeline, informed by additional resources, is essential to provide a more comprehensive and reliable support system for parents, children, and healthcare professionals. This updated timeline should be subjected to rigorous review and subsequently published in a medical context to effectively communicate the impact of screen time on neck pain across different age groups.

Doing so can guide parents, children, and healthcare professionals to make well-informed decisions about screen usage and its potential effects on neck health. This resource has the potential to significantly enhance awareness and understanding of the importance of balancing the digital world and physical well-being. It promotes the adoption of healthier habits and may contribute to reducing the prevalence of neck pain in the contemporary digital age.

## Supporting information

PRISMA Chart 1

## Data Availability

All data produced in the present work is available upon reasonable request to the authors.

## Resources

1. Kazeminasab, S., Nejadghaderi, S. A., Amiri, P., Pourfathi, H., Araj-Khodaei, M., Sullman, M. J. M., Kolahi, A. A., & Safiri, S. (2022). Neck pain: global epidemiology, trends and risk factors. BMC musculoskeletal disorders, 23(1), 26. 10.1186/s12891-021-04957-4

2. Lötters, F., & Burdorf, A. (2006). Prognostic factors for duration of sickness absence due to musculoskeletal disorders. The Clinical journal of pain, 22(2), 212–221. 10.1097/01.ajp.0000154047.30155.72

3. Global Burden of Disease Study 2013 Collaborators (2015). Global, regional, and national incidence, prevalence, and years lived with disability for 301 acute and chronic diseases and injuries in 188 countries, 1990-2013: a systematic analysis for the Global Burden of Disease Study 2013. Lancet (London, England), 386(9995), 743–800. 10.1016/S0140-6736(15)60692-4

4. Oliveira, A. C., & Silva, A. G. (2016). Neck muscle endurance and head posture: A comparison between adolescents with and without neck pain. Manual therapy, 22, 62–67. 10.1016/j.math.2015.10.002

5. Gheysvandi, E., Dianat, I., Heidarimoghadam, R., Tapak, L., Karimi-Shahanjarini, A., & Rezapur-Shahkolai, F. (2019). Neck and shoulder pain among elementary school students: prevalence and its risk factors. BMC public health, 19(1), 1299. 10.1186/s12889-019-7706-0

6. Fares, J., Fares, M. Y., & Fares, Y. (2017). Musculoskeletal neck pain in children and adolescents: Risk factors and complications. Surgical neurology international, 8, 72. 10.4103/sni.sni_445_16

7. Brattberg G. (2004). Do pain problems in young school children persist into early adulthood? A 13-year follow-up. European journal of pain (London, England), 8(3), 187–199. 10.1016/j.ejpain.2003.08.001

8. Hestbaek, L., Leboeuf-Yde, C., Kyvik, K. O., & Manniche, C. (2006). The course of low back pain from adolescence to adulthood: eight-year follow-up of 9600 twins. Spine, 31(4), 468–472. 10.1097/01.brs.0000199958.04073.d9

9. Kamper, S. J., Henschke, N., Hestbaek, L., Dunn, K. M., & Williams, C. M. (2016). Musculoskeletal pain in children and adolescents. Brazilian journal of physical therapy, 20(3), 275–284. 10.1590/bjpt-rbf.2014.0149

10. David, D., Giannini, C., Chiarelli, F., & Mohn, A. (2021). Text Neck Syndrome in Children and Adolescents. International journal of environmental research and public health, 18(4), 1565. 10.3390/ijerph18041565

11. In, T. S., Jung, J. H., Jung, K. S., & Cho, H. Y. (2021). Spinal and Pelvic Alignment of Sitting Posture Associated with Smartphone Use in Adolescents with Low Back Pain. International journal of environmental research and public health, 18(16), 8369. 10.3390/ijerph18168369

12. Brink, Y., & Louw, Q. A. (2013). A systematic review of the relationship between sitting and upper quadrant musculoskeletal pain in children and adolescents. Manual therapy, 18(4), 281–288. 10.1016/j.math.2012.11.003

13. Al-Hadidi, F., Bsisu, I., AlRyalat, S. A., Al-Zu’bi, B., Bsisu, R., Hamdan, M., Kanaan, T., Yasin, M., & Samarah, O. (2019). Association between mobile phone use and neck pain in university students: A cross-sectional study using numeric rating scale for evaluation of neck pain. PloS one, 14(5), e0217231. 10.1371/journal.pone.0217231

14. Alsalameh, A. M., Harisi, M. J., Alduayji, M. A., Almutham, A. A., & Mahmood, F. M. (2019). Evaluating the relationship between smartphone addiction/overuse and musculoskeletal pain among medical students at Qassim University. Journal of family medicine and primary care, 8(9), 2953–2959. 10.4103/jfmpc.jfmpc_665_19

15. Dianat, I., Alipour, A., & Asgari Jafarabadi, M. (2018). Risk factors for neck and shoulder pain among schoolchildren and adolescents. Journal of paediatrics and child health, 54(1), 20–27. 10.1111/jpc.13657

16. Pirnes, K. P., Kallio, J., Hakonen, H., Hautala, A., Häkkinen, A. H., & Tammelin, T. (2022). Physical activity, screen time and the incidence of neck and shoulder pain in school-aged children. Scientific reports, 12(1), 10635. 10.1038/s41598-022-14612-0

17. Page, M. J., McKenzie, J. E., Bossuyt, P. M., Boutron, I., Hoffmann, T. C., Mulrow, C. D., Shamseer, L., Tetzlaff, J. M., Akl, E. A., Brennan, S. E., Chou, R., Glanville, J., Grimshaw, J. M., Hróbjartsson, A., Lalu, M. M., Li, T., Loder, E. W., Mayo-Wilson, E., McDonald, S., McGuinness, L. A., … Moher, D. (2021). The PRISMA 2020 statement: an updated guideline for reporting systematic reviews. BMJ (Clinical research ed.), 372, n71. 10.1136/bmj.n71

18. Damasceno, G.M., Ferreira, A.S., Nogueira, L.A.C. et al. Text neck and neck pain in 18–21-year-old young adults. Eur Spine J 27, 1249–1254 (2018). 10.1007/s00586-017-5444-5

19. Vitta, A., Bento, T. P. F., Perrucini, P. O., Felippe, L. A., Poli-Frederico, R. C., & Borghi, S. M. (2021). Neck pain and associated factors in a sample of high school students in the city of Bauru, São Paulo, Brazil: cross-sectional study. Sao Paulo medical journal = Revista paulista de medicina, 139(1), 38–45. 10.1590/1516-3180.2020.0168.R1.30102020

20. Minghelli B. (2020). Musculoskeletal spine pain in adolescents: Epidemiology of non-specific neck and low back pain and risk factors. Journal of orthopaedic science : official journal of the Japanese Orthopaedic Association, 25(5), 776–780. 10.1016/j.jos.2019.10.008

21. Silva, A. G., Sa-Couto, P., Queirós, A., Neto, M., & Rocha, N. P. (2017). Pain, pain intensity and pain disability in high school students are differently associated with physical activity, screening hours and sleep. BMC musculoskeletal disorders, 18(1), 194. 10.1186/s12891-017-1557-6

22. Namwongsa, S., Puntumetakul, R., Neubert, M. S., & Boucaut, R. (2018). Factors associated with neck disorders among university student smartphone users. Work (Reading, Mass.), 61(3), 367–378. 10.3233/WOR-182819

23. Hedderson, M. M., Bekelman, T. A., Li, M., Knapp, E. A., Palmore, M., Dong, Y., Elliott, A. J., Friedman, C., Galarce, M., Gilbert-Diamond, D., Glueck, D., Hockett, C. W., Lucchini, M., McDonald, J., Sauder, K., Zhu, Y., Karagas, M. R., Dabelea, D., Ferrara, A., & Environmental Influences on Child Health Outcomes Program (2023). Trends in Screen Time Use Among Children During the COVID-19 Pandemic, July 2019 Through August 2021. JAMA network open, 6(2), e2256157. 10.1001/jamanetworkopen.2022.56157

24. Ayhualem, S., Alamer, A., Dabi, S. D., Bogale, K. G., Abebe, A. B., & Chala, M. B. (2021). Burden of neck pain and associated factors among smart phone user students in University of Gondar, Ethiopia. PloS one, 16(9), e0256794. 10.1371/journal.pone.0256794

25. Al-Hadidi, F., Bsisu, I., AlRyalat, S. A., Al-Zu’bi, B., Bsisu, R., Hamdan, M., Kanaan, T., Yasin, M., & Samarah, O. (2019). Association between mobile phone use and neck pain in university students: A cross-sectional study using numeric rating scale for evaluation of neck pain. PloS one, 14(5), e0217231. 10.1371/journal.pone.0217231

26. Gheysvandi, E., Dianat, I., Heidarimoghadam, R. et al. Neck and shoulder pain among elementary school students: prevalence and its risk factors. BMC Public Health 19, 1299 (2019). 10.1186/s12889-019-7706-0

27. Shan, Z., Deng, G., Li, J., Li, Y., Zhang, Y., & Zhao, Q. (2013). Correlational analysis of neck/shoulder pain and low back pain with the use of digital products, physical activity and psychological status among adolescents in Shanghai. PloS one, 8(10), e78109. 10.1371/journal.pone.0078109

28. Buabbas, A.J., Al-Mass, M.A., Al-Tawari, B.A. et al. The detrimental impacts of smart technology device overuse among school students in Kuwait: a cross-sectional survey. BMC Pediatr 20, 524 (2020). 10.1186/s12887-020-02417-x

29. Cankurtaran, F., Menevşe, O., Namlı, A., Kızıltoprak, H. Ş., Altay, S., Duran, M., Demir, E. B., Şahan, A. A., & Ekşi, C. (2022). The impact of digital game addiction on musculoskeletal system of secondary school children. Nigerian journal of clinical practice, 25(2), 153–159. 10.4103/njcp.njcp_177_20

30. Fares, J., Fares, M. Y., & Fares, Y. (2017). Musculoskeletal neck pain in children and adolescents: Risk factors and complications. Surgical neurology international, 8, 72. 10.4103/sni.sni_445_16

31. Yang, S. Y., Chen, M. D., Huang, Y. C., Lin, C. Y., & Chang, J. H. (2017). Association Between Smartphone Use and Musculoskeletal Discomfort in Adolescent Students. Journal of community health, 42(3), 423–430. 10.1007/s10900-016-0271-x

32. Crawford, J. O. (2007). The Nordic Musculoskeletal Questionnaire. Occupational Medicine, 57(4), 300–301. 10.1093/occmed/kqm036

33. Bahkir, F. A., & Grandee, S. S. (2020). Impact of the COVID-19 lockdown on digital device-related ocular health. Indian journal of ophthalmology, 68(11), 2378–2383. 10.4103/ijo.IJO_2306_20

34. CDC. (2018, January 29). Infographics - Screen Time vs. Lean Time | DNPAO | CDC. Centers for Disease Control and Prevention. Retrieved March 13, 2024, from https://www.cdc.gov/nccdphp/dnpao/multimedia/infographics/getmoving.html

35. Robinson, T. N., Banda, J. A., Hale, L., Lu, A. S., Fleming-Milici, F., Calvert, S. L., & Wartella, E. (2017). Screen Media Exposure and Obesity in Children and Adolescents. Pediatrics, 140(Suppl 2), S97–S101. 10.1542/peds.2016-1758K

36. Farhangi, M.A., Fathi Azar, E., Manzouri, A. et al. Prolonged screen watching behavior is associated with high blood pressure among children and adolescents: a systematic review and dose–response meta-analysis. J Health Popul Nutr 42, 89 (2023). 10.1186/s41043-023-00437-8

37. Chu, G. C. H., Chan, L. Y. L., Do, C. W., Tse, A. C. Y., Cheung, T., Szeto, G. P. Y., So, B. C. L., Lee, R. L. T., & Lee, P. H. (2023). Association between time spent on smartphones and digital eye strain: A 1-year prospective observational study among Hong Kong children and adolescents. Environmental science and pollution research international, 30(20), 58428–58435. 10.1007/s11356-023-26258-0

38. Maurya, C., Muhammad, T., Maurya, P. et al. The association of smartphone screen time with sleep problems among adolescents and young adults: cross-sectional findings from India. BMC Public Health 22, 1686 (2022). 10.1186/s12889-022-14076-x

39. Blozik, E., Laptinskaya, D., Herrmann-Lingen, C., Schaefer, H., Kochen, M. M., Himmel, W., & Scherer, M. (2009). Depression and anxiety as major determinants of neck pain: a cross-sectional study in general practice. BMC musculoskeletal disorders, 10, 13. 10.1186/1471-2474-10-13

