## Supplementary material for "Screen Time and Musculoskeletal Neck Pain in Children: A Comprehensive Systematic Review and Lifestyle Recommendations": PRISMA Chart 1

**PRISMA 2020 flow diagram for new systematic reviews which included searches of databases and registers only**

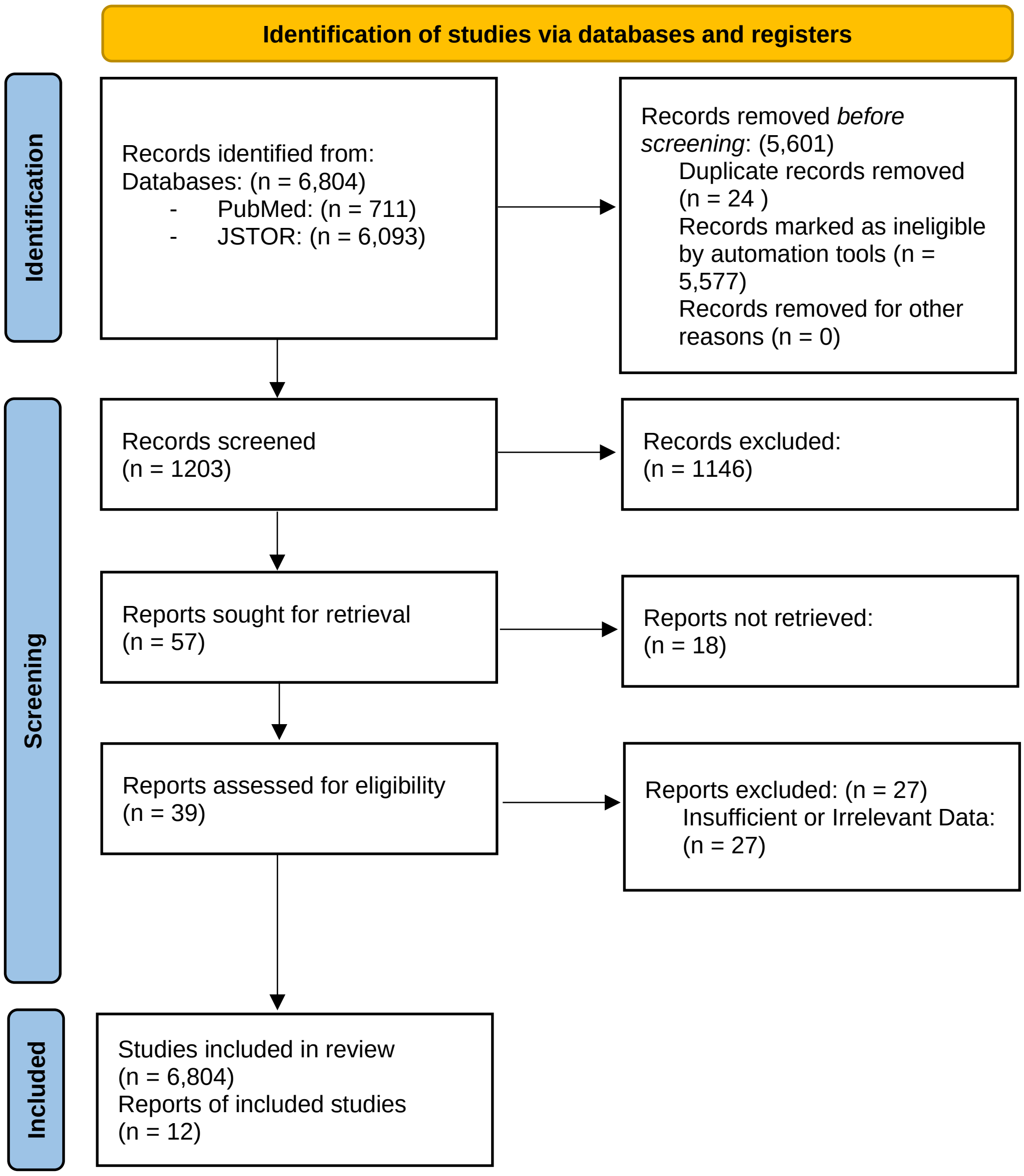
