## Supplemental Files 2 for "Screen Time and Musculoskeletal Neck Pain in Children: A Comprehensive Systematic Review and Lifestyle Recommendations"

**Supplemental Files 1**

***PubMed Search String 1***

*((technology) OR (smartphone) OR (cell phone) OR (television) OR (screen time) OR (text)) AND ((postural habits) OR (posture) OR (neck pain) OR (lower back pain)) AND ((adolescence) OR (children) OR (youth) OR (babies) OR (toddler))*

2,328 results (before filters)

***JSTOR Search String***

*((neck pain) OR (neck posture) OR (cerviclegia)) AND ((adolescence) OR (youth) OR (children) OR (young adult)) AND ((technology) OR (cell phone) OR (phone))*

8,802 results (before filters)
